# Extrication of patients trapped following a motor vehicle collision: a systematic scoping review of the literature

**DOI:** 10.1101/2024.06.10.24308701

**Authors:** Tim Nutbeam

## Abstract

**Background:** Extrication is the process of removing injured or potentially injured people from their vehicles. The origin of current extrication techniques and paradigms is largely unknown. An understanding of the historical evidence related to motor vehicle collisions (MVCs), injuries and deaths will provide context for accepted, contemporary, extrication practices.

**Methods:** Extrication related search terms were developed and applied across of range of sources including Clinical and health care data, Trial registries, Grey literature, Academic and specific Transport related sources.

**Results:** 7089 articles were identified, following review, 170 are included in this qualitative synthesis. Key themes / categories included: Extrication training and principles, Injures, Immobilisation, Care during entrapment, Clinical response type, Vehicle deformity intrusion entrapment, and Extrication.

**Conclusion:** There is a paucity of published evidence to support the current approach to extrication of entrapped patients following an MVC. Focused studies identifying in detail the injures and their sequelae associated with entrapment, the biomechanics of current techniques and ensuring that the patient perspective is captured will enable the development of much needed evidence based multidisciplinary guidance.

## Background

Extrication is the process by which injured, or potentially injured casualties are removed from their vehicles following a motor vehicle collision (MVC) [1]. The origin of current extrication techniques and paradigms is largely unknown. An understanding of the historical evidence related to MVCs, injuries and deaths will provide context for accepted, contemporary, extrication practices.

The review objectives can be defined by the following research questions [2]:

- What is the (historical and scientific) context for current extrication approaches as delivered by rescue services?
- What injuries are sustained by patients who are trapped in their motor vehicles and how does this influence extrication practice?
- What are the needs of patients who are trapped following an MVC, how are these met and following extrication where is their care best delivered?

Extrication is a multidisciplinary undertaking; the literature originates from a wide range of disciplines (clinical, rescue, vehicle design and testing). A systematic scoping approach was considered most appropriate for this review due to both the predicted heterogeneity of the literature and the overarching purpose of this review: to identify gaps in the literature which will aid in the planning of future research [3]. This review will describe and give context to the evolution of the current operational and clinical approach to extrication and identify areas where additional knowledge should be prioritised.

For the purposes of this review, extrication is considered as “the rescue and removal of patients from motor vehicles following a collision”. This review does not include other specialist areas such as rescues from water, caves or collapsed buildings. This review excludes the technical detail of rescue practice and the details of specific rescue equipment. This scoping review is reported to Preferred Reporting Items for Systematic reviews and Meta-Analyses extension for Scoping Reviews (PRISMA-ScR) guidance [4].

## Methods

### Search strategy

This is a scoping review; papers and sources were identified through a systematic search strategy based upon PRISMA methodology. The aim was not to identify a single three-part question – but to identify literature that would inform a deep understanding extrication and associated themes (see question statements above).

### Development of search terms

The search strategy was developed with professional librarian assistance, trialled, and further refined to ensure that appropriate references and sources were not missed. The final search strategy is summarised in the box below.

#### Box 1. 1 Search Terms used

1. Extrication OR immobilisation OR intrusion OR roof removal OR side rip OR self-extrication OR chain cabling
2. Car OR motor vehicle OR automobile OR vehicle OR road Traffic OR accidents OR traffic OR collision
3. (MVC OR MVA OR RTA OR RTC) and (collision OR accident)

Search: (i OR ii) AND (iii OR iv)

The following were searched in August 2021:

Clinical and health care data sources: National Health Services (NHS) available databases using the Healthcare Databases Advanced Search function which includes Medline, EMBASE, CINAHL, EMCare, Healthcare Management Information Consortium (HMIC). From the Cochrane Library we searched the Cochrane Database of Systematic Reviews, Cochrane Central Register of Controlled Trials and Cochrane Clinical Answers. In addition, we searched the Web of Science, Scopus, Health Foundation, Nuffield Trust, PLOS ONE, TRIP, and the Knowledge for Health Care databases.

Trial registries: Clinictrials.gov, WHO International Clinical Trials Registry Platform, EU clinical trials register and the International Standard Randomised Controlled Trial Number ISRCTN registry.

Grey literature sources: The National Grey Literature Collection via the MEDNAR interface, The OAIster^®^ database, The CORE repository, Open Grey, Grey Matters.

Academic sources: E-theses online service (EThOs) from the British Library, Networked Digital Library of Theses and Dissertation (NDLTD), Open Access Theses and Dissertations (OATD)

Other data sources: safetylit.org, the international transport forum web interface, the national academic of science engineering and medicine and the international research council on biomechanics of injury.

### Selection of studies

Following the search, the Endnote interface (EndNote X9.3.3, Clarivate, Philadelphia, PA, 2013) was used to identify and remove duplicate articles. Sources were included for further review which were and available in the English language and available online or through library services. The remaining studies were reviewed using their abstract and studies which were not relevant to the research questions excluded. A full-text review allowed further exclusion of articles that were not relevant to the research question. Remaining articles were included and their reference list reviewed to identify further articles for inclusion.

### Synthesis

Following exclusions, full text sources were reviewed with reference to the research questions and a broad analysis of the domains identified conducted; articles were grouped into domains, reviewed and included in the narrative discussion.

## Results

An initial total of 16,413 documents were identified through the search strategy. This was reduced to 7089 following removal of duplicates. One hundred and seventy papers were identified that were relevant to the research questions. Results are summarised in Figures 1.1 and 1.2.

**Figure 1.1:**
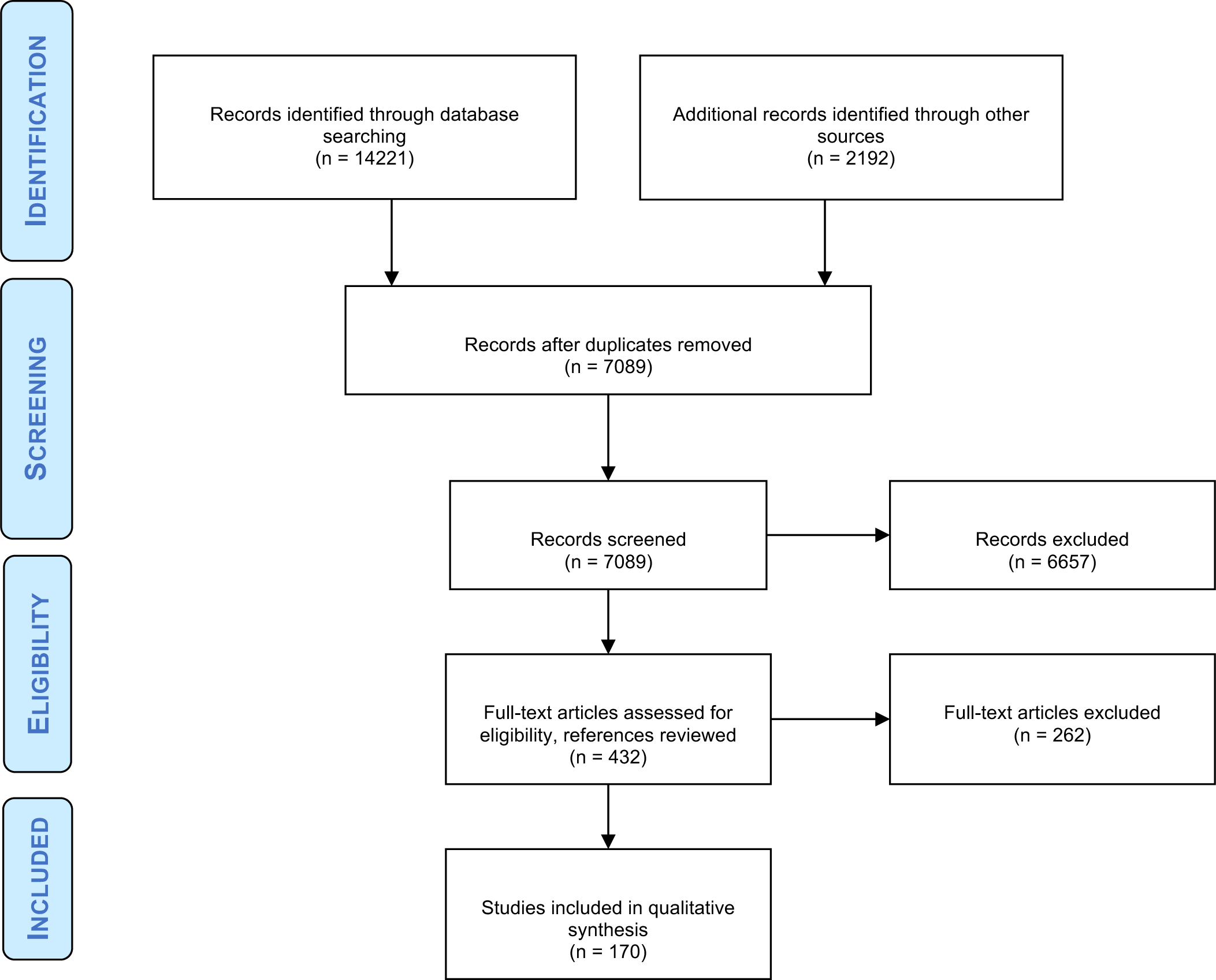
Studies screened and included (adapted from PRISMA)

**Figure 1.2.**
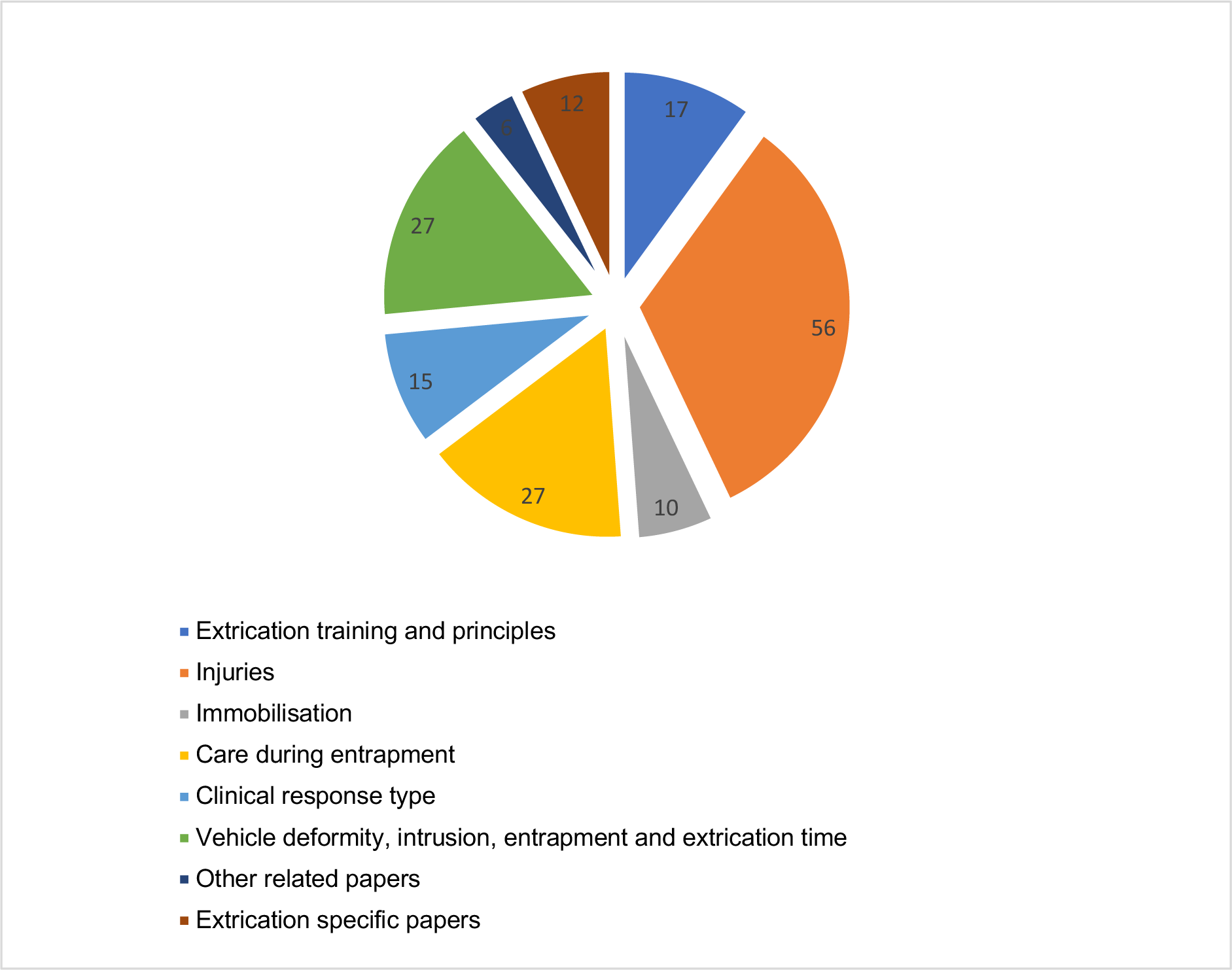
Domains identified.

Figure 1.2 Outlines the domains which were identified by full-text review. These are: extrication training and principles, injuries associated with MVC and extrication, immobilisation, care during entrapment, clinical response type and vehicle deformity, intrusion, entrapment and extrication time, other related papers and extrication specific papers.

## Discussion

The scoping review identified studies from a wide range of disciplines and backgrounds. The study types were diverse including computer modelling of accidents and energy transfer, retrospective chart review and database review studies, individual case reports, case series, post-mortem studies, biomechanics, kinematics and mannequin-based studies. There were no randomised controlled trials, no interventional studies of clinical or operational care, and no prospective cohort studies. There were only two unique prospective ‘real world’ extrication focused analyses [5,6].

Common domains in the literature are explored in the following sections.

### Extrication training and principles

The principle of movement minimisation is a key paradigm of contemporary extrication practice [1]. The earliest papers that discussed the priorities to achieve in extrication are from medical journals in the 60’s and 70’s. These papers identify that patients may have time dependent injuries and state the importance of movement minimisation to prevent avoidable secondary spinal injury following an MVC [7–13]. The assertions in relation to movement minimisation are made without reference to specific cases, case series or published data. The primacy of movement minimisation during the process of extrication emerges in extrication manuals and guidance aimed at rescue services from the 1970’s and onwards [14–22]. The manuals and textbooks were unreferenced in respect to the origin of, or justification for the primary focus on spinal injuries above other injuries in the development of extrication paradigms.

### Injuries

Early post-mortem studies identify the wide range of injuries from which patients injured in MVC succumbed [23]. Even in these early studies (and before the introduction of modern safety systems) the rate of spinal cord injury and particularly isolated spinal injury (which might justify movement minimisation extrication methods for extrication) was low compared to other injuries; 0.8% of fatalities had a spinal cord injury and 70% had a head injury [23]. With the adoption of seatbelts, the primary injuries and death caused by ejection were minimised with an associated drop in mortality, but new injuries originated: facial fractures from impacting with the internal surfaces of the car and abdominal injuries cause by the belts themselves [24,25].

Much of the literature focuses on injuries in isolation, as opposed to patterns or constellations of injuries. Several papers consider individual cases, case series and mechanism type for a variety of individual injuries including limb [26–29], aortic [30–34], pelvic [35–37] brain [38–41], abdominal [30,42,43] and other body areas and injury types [44–50]. Injured obese patients were identified as having worse outcomes [51].

### Spinal injuries

Case reports and retrospective reviews of routinely collected data of severe spinal injury following an MVC featured both adults and children [52–56]. Mezue *et al.* reported failures in prehospital immobilisation and careful handling in patients with subsequently proven spinal cord injury [57]. The authors report that 94.1% of patients in their series were extricated by bystanders and only 36% of the patients had any attempt at immobilisation prior to hospital arrival, the authors report an association between adequate immobilisation and transport and improved function at discharge (p=0.003) [57].

Sochor and colleagues identify scene factors which predict the presence of a clinically important spinal injury [58]. In front seat restrained drivers or passengers between 16 and 60 years of age, if the glass in their car was unbroken following an MVC that the rate of clinically important spinal injury was very low. The sensitivity for the GLASS rule was 95.20% (95% CI 91.45–98.95%), specificity was 54.27% (95% CI 53.44–55.09%), and the negative predictive value was 99.92% (95% CI 99.86–99.98%) [58].

### Injuries in those who are trapped

Siegel *et al.* compared injuries in patients who required extrication compared to those that did not. They found a higher rate of brain (51% v’s 35%, non-significant), lower extremity (58% v’s 30%, p<0.003) and splenic injuries (22%, v’s 10%, p<0.02) in patients that required extrication compared to those that did not [59].

Sanson *et al.* report a case series of HEMS delivered critical care interventions on patients who were trapped. They report a high injury load including tension pneumothorax (11.8%), major head injury (39%), and non-compressible haemorrhage (34.7%) [60]. Wilmink reports a case series of entrapment MVCs attended by a UK Helicopter Emergency Medical Services (HEMS) [6], with a high injury load (median ISS 17, range 1-59) in entrapped patients and an associated high mortality (10%). They note that in their case series isolated spinal cord injury did not occur with a majority of patients with severe spinal injury having an associated major head injury affecting their level of consciousness and therefore limiting the efficacy of clinical assessment (36% of all patients had a head or spinal injury) [6]. Westhoff *et al.* consider trapped patients from both passenger vehicles and trucks and report a high degree of severe single system injury (68.7% to the head, 23.5% to the neck, 50.8% to the chest, 43.6% to upper extremities, 15.4% to the abdomen, 16.4% to the pelvis, and 52.9% to lower extremities) and multiple injuries in trapped patients [61].

The literature identified in this scoping review does not provide contemporaneous data that allows us to accurately report the rate of spinal cord injury in entrapped patients. We can conclude that the rate of time dependent injury is high in the those who are entrapped but it is unclear if this is leads to poor outcomes or if entrapment alone might lead to increased morbidity and mortality.

### Non-physical injuries

Non-physical injuries are a frequent cause of long-term morbidity and affect the quality of life of those who suffer from them [62]. MVC’s are associated with a high rate of post-traumatic stress disorder (PTSD) and other psychological sequelae both in children and adults [63–78]. Mayou’s group compared those with multiple injuries following an MVC and those with whiplash injuries alone, they report that in the acute phase (within one month) following the accident that those with multiple injuries were more likely to have an acute stress reaction (41%, comparator not reported); interestingly long-term psychological outcomes did not appear to be correlated with severity of injury [65]. Mosaku K *et al*. performed a complimentary study that identified that clinical factors did not predict long term psychological outcomes [77]. Heron-Delaney conducted a systematic review with the intention of identifying factors that predict PTSD in adult MVC survivors and found that the prevalence of PTSD varied from 6-45% with a “perceived threat to life” being a significant predictor of long term poor psychological recovery [79]. Watts and team found that up to 77% of post MVC victims admitted to hospital were likely to have an “acute psychiatric disorder” with 11-15% seeking or receiving professional counselling [69].

Arnberg and team considered the long-term PTSD outcomes of children following an MVC; they found a high prevalence of stress reactions at nine months following the event (50-69%), with PTSD symptoms still present in 18% of their sample at 20 years [73].

A single paper considered the experience of patients that were trapped (due to spinal cord injury) following an MVC [80]. Sepahvand introduces the concept of “scene shock” in which the injured and untrained bystanders fall into a “state of instability” leading to emotionally driven decisions and subsequent behaviours that lead to desperate, unplanned rescue efforts which may contribute to secondary spinal cord injury [80].

This review confirms that non-physical injuries are common following MVC. Specific data on entrapment or extrication as a risk factor for non-physical injury was not identified. We hypothesise that being trapped would be considered by patients to be a “threat to life” and as such this group may be at higher risk of poor psychological outcomes and long-term symptoms. Importantly, no data was available that recounted the patient experience of entrapment or extrication or considered if changes to this area of practice may improve the patient experience.

### Immobilisation

Prehospital services use immobilisation devices to mitigate against movement and ensure or return anatomical normality [81]. Immobilisation can include the application of a femoral traction device, a pelvic sling or the ‘triple immobilisation’ of a cervical collar, head blocks and a long board or scoop stretcher. Two papers in this review reported pelvic immobilisation techniques and suggest that they may be appropriate for use in entrapment [82,83]. A small number of papers reported methods of paediatric immobilisation using novel techniques or adapting standard prehospital equipment [84–86].

Recent publications challenge the ubiquitous application of cervical collars or the use of spinal boards in the extrication and transportation phase following an MVC [87–90]. These papers, based on expert opinion and an analysis of ‘excess imaging’ associated with immobilisation suggest alternative approaches including gentle patient handling techniques and self-extrication [87–90].

Immobilisation, particularly the use of cervical collars has been a subject of increasing enquiry and consideration over the last 15 years [91–93]. Authors have challenged the harm / benefit of collar application, particularly in conscious trauma patients [93]. The use of such immobilisation devices specifically in the context of entrapment and extrication is discussed later in this review.

### Care during entrapment

Papers were identified that related to the delivery of patient care, minimisation of patient harm or improvement of patient experience during entrapment. No papers were identified which included any description of patient experience or collection of patient generated data (e.g. pain scores).

Single case studies were presented which identify pain and the potential for hypothermia as issues that benefit from mitigation whilst the patient remains trapped [94–97]. A series of four cases supported by a literature review identify that ketamine is well suited for meeting the analgesic needs of a trapped patient [98]. Further papers presented general principles and opinion on pain management options [99,100].

A surprisingly large number of mannequin-based studies evaluated the use of a wide variety of laryngoscopes or supraglottic airway devices for the placement of endotracheal tubes in entrapped mannequins in various positions [101–115]. Individual case studies and small case series supported the use of supraglottic airway devices in extremis [116–118]. A single retrospective chart review of airway management published as an abstract recognised the challenges of intubation in the entrapped patient [119].

The literature in this area is limited to a single case series, expert opinion and mannequin studies looking exclusively at airway management. Literature was not identified that defined patient’s clinical needs and priorities for the management during the entrapment and extrication phase.

### Clinical response type

The utility of bystanders at the scene of an MVC was considered by several authors. Thierbach *et al.* identified that bystanders were more likely to help with those with moderate injuries than patients with severe injuries and advocated for more advanced widely available bystander training [120]. Heightman and Bhalla discuss the potential utility of bystander care to reduce mortality and morbidity, especially with those with specific training, kit and authorisation [121,122]. Bhalla reflects on the potential medico-legal culpability for bystanders in providing immediate care and how this might be overcome by training and authorisation to act [121,122].

Two studies from the 1990’s identified that entrapment was associated with severe injuries, and this resulted in complex patient care needs which were often unmet [123,124]. Many papers advocated for physician attendance at scene for entrapment trauma [125–131]. A single prospective cohort study considering all mechanisms of major trauma found no survival benefit when a physician was present (OR of 1.16 (95% confidence interval = 0.97 to 1.40, p = 0.11).

Byrne *et al.* report that longer response times were associated with higher rates of mortality [132], whilst Gauss and team noted the association between prolonged prehospital time and poor patient outcomes [133].

Patients who are trapped have on average longer prehospital timelines and as such may have an excess mortality for this reason alone [134]. The benefits and potential harms of bystanders to patients and the ideal clinical response model cannot be surmised from the literature available to this review.

### Vehicle deformity, intrusion, entrapment and extrication time

These papers considered patient and incident-based factors which predicted (or failed to predict) mortality, injury or the need for trauma centre care. The papers offered different perspectives as to the utility of incident-based factors both in isolation and combined with injury, physiological or patient demographic factors.

The factors of interest to this review are the association between vehicle structural deformity (external), intrusion into the passenger compartment and the requirement for the extrication of a patient. These factors are important to our question of the inter-relation of patient injury and their ability to self-extricate.

Three papers considered the accuracy of the data recorded by both paramedics and emergency physicians in terms of scene characteristics (such as need for extrication). Poor completion of prehospital records and poor correlation between findings at scene and subsequent analysis led to both under and over triage (EMS record accuracy median 28.5%, range 0-100%) [135–137].

### Deformity

External vehicle deformity was found to be important when combined with intrusion in the absence of air bags (OR 5.2, 95% CI 2.525–10.780) [138]. Deformity was also important in predicting mortality in older patients (differences in mortality were associated with age (OR 6.92,95% CI 1.2-38.9) and a high vehicle deformity (OR 3.28, 95%C1 1.5-6.8)[139].

Intrusion:

Studies reached different conclusions when considering intrusion alone as a predictor of injury, mortality or trauma system utilisation [140,141]. One paper identified supported the utilisation of intrusion alone in frontal collisions as an indicator of major trauma and as such should feature on major trauma triage tools [142]. A paper from 1996 reported the utility of intrusion of >24 inches as a triage criterion but found other mechanistic criteria were not useful [143]. Davidson *et al.* found that intrusion of more than 12 inches were useful in predicting trauma centre utilisation over and above physiological criteria; they found mechanistic criteria particularly useful in older patients without physiological derangement. Intrusion of greater than 12 inches had a PPV of 10.4% (95% CI, 9.5-11.3) to predict severe injury; steering wheel collapse had a PPV of 25.7% (95% CI, 23.0-28.4%) for the same outcome [144].

More recent reviews did not support intrusion as a stand-alone predictor of injury, and instead suggest that patients triaged on intrusion alone had low Injury Severity Score (ISS) and a high discharge rate (ISS was 5 (1.75, 10.25) and 39.5% were discharged from the Emergency Department (ED)) [145,146]. Simon *et al.* recommend that if certain mechanistic features were present and no evidence of physiological disturbance then an initial clinical review of the patient should occur and then upgrade to a trauma team if required [147]

The combination of intrusion and entrapment, which are often inter-related, was identified as useful for predicting patient mortality. When adjusted for age and sex, the following mechanism of injury (MOI) were associated with mortality: passenger space intrusion (OR 1.74; CI 1.18, 2.57), extrication (OR 2.16, CI 1.14, 4.04), ejection (OR 8.33; CI 4.68, 14.83) and occupant fatality (OR 2.28; CI 0.50, 10.40) [148].

### Entrapment and extrication

Many groups identified that entrapment, particularly when associated with prolonged or difficult extrication (typically defined as > 20 minutes) was a useful predictor of injury (multivariate OR 2.5, 1.1–6.0, p=0.04), and was a more sensitive and specific criterion for trauma centre utilisation than other mechanistic features [149–156].

This finding was not universal with two authors recommending that the need for extrication in isolation should be removed from triage guidance as it led to considerable over-triage [157,158].

There were no studies concerning vehicle deformity or extrication which included children. However, intrusion was found to be associated with increased injury in children, with a direct relationship between the amount of intrusion and associated injuries (4.0% increase in AIS3+ injuries for each cm of intrusion (95% CI = 2.7-5.2%) [159–161].

### Other related papers

Ryb *et al.* suggested that patient mobility post collision was more useful than mechanistic factors in triaging patients to an appropriate facility; self-extrication under-triaged by 0.4% as a predictor of death[162]. Schulman and colleagues developed a composite “Scenescore” consisting of weighted values for age, collision type, impact location, airbag deployment, steering wheel deformity, intrusion, and restraint use; they suggest a score of 8 offers optimal performance (sensitivity 76%, specificity 46%) to assist with triage decisions [163]. Technological solutions were also suggested utilising automatic crash notification or vehicle telemetry to predict injuries and inform response [164–166].

As might be expected the conclusions and recommendations varied with the era of analysis and publication. This may be in part to the increased safety of vehicle systems, the development of vehicles in terms of crumple zones, changes in the way patients were considered trapped or needed extrication and the individual capability and acceptable over-triage rates of the system under consideration.

### Extrication specific papers

Nutbeam *et al.* prospectively collected data at the scene of entrapment MVC, then used this to report factors that predict the need for extrication, the factors which affect this time and the number of extrications in which physical or actual entrapment occurs (10%) [5,134,167]. This low rate of physical entrapment (10% of all extrications), the time taken for extrication (median 30 minutes) and the increased mortality seen with both entrapment and increasing time between injury and arrival at hospital demonstrates the importance of the entrapped patient as an area where increased knowledge and decreasing the rate and time of entrapment may lead to improved patient outcomes.

There were very few papers that considered the effect of extrication technique on entrapment time or patient outcomes. Lars and Fattah both demonstrate the speed of chain cabling type techniques which are used in Scandinavian countries but not frequently used elsewhere compared to more traditional techniques in experimental conditions [168,169].

There are a number of papers that report bio-mechanical analysis using various methodologies and a range of extrication types. Bucher *et al.* found that utilising a KED (Kendrick extrication device) resulted in less spinal movement in patients with a normal body mass index (BMI) but increased spinal movement in obese patients [170]. Shafer *et al.* performed a pilot study which concluded that allowing an individual to exit a car under their own volition (self-extrication) with a cervical collar in place may result in the least amount of motion compared to exiting with paramedic assistance [171]. These findings were reinforced by Engsverg, Gabrieli, Haske and Dixon and their respective teams across a number of extrication methods using a variety of biomechanical methods and outcome measures [172–176].

### Where are the gaps?

Considering the large number of patients whose clinical care, timeline to hospital and patient experience may have been adversely affected by their trapped status, there is little focused literature which allows an understanding of key areas of this phenomenon which would enable an EBM approach to the development of evidence-based extrication guidance.

**Figure 1.3.**
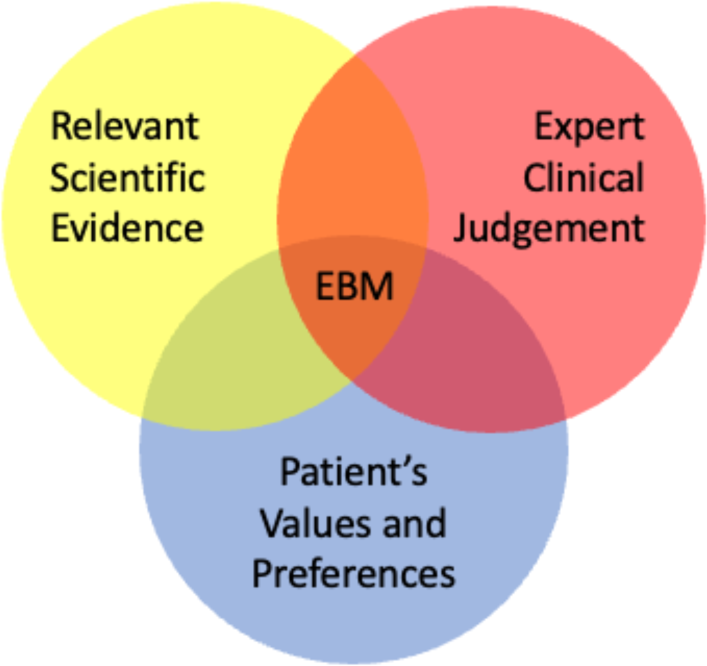
The EBM Triad.

Areas of ‘relevant scientific evidence’ where data is not available or not sufficient includes the difference in injury patterns between trapped and not trapped patients, the difference in outcome between trapped and not trapped patients, the efficacy of extrication techniques to minimise movement and their clinical or outcome implications. There is not currently evidence that enables us to understand ‘patient values and preferences’; we do not have data which supports an understanding of the patient experience of extrication and how this may be improved. Despite a large number of case reports and papers from single or small groups of experts there is no coherent, consensus “expert clinical judgement’ which bridges the rescuer-clinician divide in the current literature. The absence of multidisciplinary guidance based on the best available evidence demonstrates another notable gap in relation to this important patient group.

Our understanding of these important areas of research could be improved by targeted studies analysing high-quality data sources which allow comparison of injuries, injury patterns and outcomes between trapped and not-trapped patients following an MVC. Such analyses will be enhanced by reporting the frequency of isolated spinal injuries that may be exacerbated by movement and time-critical injuries such as significant head injuries. These analyses will contextualise the risk of secondary spinal injury, the risks of patient deterioration whilst trapped and help us to understand the potential for self-extrication. Sub-analyses which allow comparisons between patients of different ages, sex and body habitus will further inform decision making in this area.

Current biomechanical data of extrications are limited to small numbers of extrications across a small pool of healthy volunteers. Where possible, real-world data should be collected to inform our understanding of the performance of currently deployed extrication techniques. If real-world data collection is not possible then researchers should deliver adequately powered studies which consider all extrication types across a range of people.

Good evidence-based medicine requires the consideration of patient values and preferences [177]. The absence of the patient voice from the current evidence base is notable and rectifying this should be a target for future research. Patient surveys and interviews will assist in capturing the patient perspective and routinely collected data in this area should include patient experience. Patient priorities should be identified, and patient representatives should be engaged in the development of guidance for the care of patients whilst they are entrapped and subsequently extricated.

Solutions for the evidence gaps identified above will enable the development of much needed evidence-based multidisciplinary guidance through consensus processes.

### Limitations of this scoping review

We aimed for a comprehensive search strategy; however, it may have missed studies that were important to our defined questions. Steps were taken to keep the inclusion criteria broad and included a large number of grey literature sources; which in turn required the review of a large number of papers. By defining questions in advance, we attempted to produce a decision-making process which was predictable and reproducible, but this was not confirmed through any verification process. A single researcher applied the questions and made decisions regarding inclusion and exclusion criteria, which may have improved the reliability of these decisions but threatens the reproducibility if repeated by another person or team.

The nature of the scoping review does not require a formal risk of bias assessment. The broad nature of the review does not allow for the comprehensive synthesis of all domains, nor does it provide the specificity to identify immediate recommendations to improve extrication practice.

## Conclusions

There is a paucity of published evidence to support the current approach to extrication of entrapped patients following an MVC. Focused studies identifying in detail the injures and their sequelae associated with entrapment, the biomechanics of current techniques and ensuring that the patient perspective is captured will enable the development of much needed evidence based multidisciplinary guidance.

## Data Availability

All data produced in the present work are contained in the manuscript.

## Acknowledgements

This systematic scoping review was submitted as chapter in The Development of Evidence-Based Guidelines to Inform the Extrication of Casualties Trapped in Motor Vehicles following a Collision, towards Doctor of Philosophy (Phd) In Emergency Medicine, University of Cape Town 2022. Supervisors: Dr Willem Stassen, Professor Jason Smith, Professor Lee Wallis

